# Characteristics and evolution of COVID-19 cases in Brazil: mathematical modeling and simulation

**DOI:** 10.1101/2020.10.14.20212829

**Authors:** Carlos Augusto Cardoso Passos, Estéfano Aparecido Vieira, José André Lourenço, Jefferson Oliveira do Nascimento

## Abstract

The pandemic caused by the coronavirus of severe acute respiratory syndrome 2 (SARS-CoV-2), the etiological agent of the 2019 coronavirus disease (COVID-19), represents a threat of great magnitude not faced in this century. As a result, each government has proposed emergency public health measures that are critical to delay the transmission and spread of the virus and mitigate its impacts. In Brazil, the outbreak triggered many cases of people infected with COVID-19. Considering there are no drugs or vaccines proven to be effective to treat the disease, analyzing the data of infection cases and their mathematical interpretation are essential for supporting and guiding governmental measures to suppress and mitigate the impact of COVID-19. This means that estimates with mathematical models to assess the development potential of sustained human-human transmission are needed. Since the disease has its own biological characteristics, the models need to be adapted to the variability of the regions’ characteristics and the decision-making by both the government and the population, in order to be able to deal with real situations. Thus, in the present paper, we analyzed the official data of COVID-19 in Brazil and used the Johnson-Mehl-Avrami-Kolmogorov (JMAK) equation to predict the evolution of the disease. The model indicates that a nucleation rate is of fourth order, which indicates that Brazilians are crowding with no respect to measures of social distance and disease prevention. In our opinion, the political authorities were unable to control the spread of the disease in Brazil, given that social mobility was interrupted by the federal and state governments.

## INTRODUCTION

Given the global scenario of the pandemic caused by the new coronavirus, the severe acute respiratory syndrome-associated coronavirus (SARS-CoV-2), each government has proposed fundamental emergency measures to halt the transmission and spread of the virus and mitigate its impacts. Specifically in Brazil, the outbreak first occurred by importing the pathogen into São Paulo State,^1^ and then into other regions of the country, which triggered a large number of cases of people infected with the new coronavirus. Soon after, on March 20^th^, 2020, the Brazilian Ministry of Health declared community-acquired infection.^2^

Considering there are no vaccines or drugs proven to be effective for the treatment of coronavirus disease (COVID-19), analyzing data of infection cases and their mathematical interpretation is important to support and guide the government measures for suppressing and mitigating the impacts of COVID-19. This means that making estimates with mathematical models is needed to assess the potential development of sustained human-to-human transmission.^3^

Because the disease has its own particular biological characteristics, models must be adapted to each specific case, to the regions’ characteristics, and to decision making by the government and the population for being capable of dealing with real situations^4,5^. In other words, this is a new virus that creates a completely new situation.

In literature, some examples of mathematical models related to the dynamics of infectious diseases can be found. Different types can be used, essentially considering non-linear equations.^6-9^ The dynamics description of coronavirus infection in humans requires the simulation of different scenarios to avoid false estimates. According to Verity et al.,^9^ during the exponential growth phase of an epidemic, the time observed between the onset of symptoms and results (recovery or death) is suppressed, and naive estimates of the times observed provide biased responses of real time. According to these authors, ignoring this effect tends to influence the down estimate in the number of deaths.^9^

In this context, with data from COVID-19 outbreaks, our research group initiated the outbreak modeling and simulation considering the social distance measures adopted in Brazil.^10^ Our main objective is to predict the cases of the new coronavirus by finding out the transmission characteristics of COVID-19 in Brazil, in specific regions, to assist in the decision-making by government authorities. To this end, we use a crystal growth model to analyze the data obtained and describe the limitations of this methodology.

## METHODOLOGY

We used the database of the Brazilian Ministry of Health to make the estimate. For analysis, we needed to obtain the approximate size of the population (N) of each state and of Brazil. In this case, we adopted the population estimates for each state calculated by the Brazilian Institute of Geography and Statistics (*Instituto Brasileiro de Geografia e Estatística* - IBGE) for 2019.^11^

Our prediction model is based on the Johnson-Mehl-Avrami-Kolmogorov (JMAK) theory. This theoretical model is widely used in Materials Science and Engineering, Computing, Economics and Ecological Systems. It specifically describes processes for transforming solid materials.^12-15^ Equation 1 is the original expression of the JMAK theory and it read as follows:

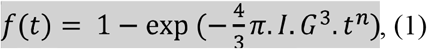

In which:

*I* = the number of nuclei or a rate of nuclei emergence; G = the growth rate of each nucleus;

*t* = time;

*n* = a parameter that represents whether the system follows site saturation. This way, the value will be equal to 3; under constant nucleation rate, the value will be equal to 4.

In the case of constant nucleation rate, the rate of nuclei emergence is described by Equation 2:

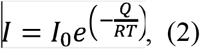

in which:

*Q* = activation energy;

*R* = ideal gas constant (*R* = 8.314 J/K.mol).

When Equation 1 is applied to phase transformation processes of materials, the constant nucleation rate must be considered under two different situations. The first is with site saturation, in which the nuclei appear for a given time t=0 and all grow at the same rate. Thus, the volume of each nucleus is the same for all nuclei until 100% of the transformation occurs. Over time, no new nuclei appear, and the term *I* in Equation 1 assumes a fixed value throughout the transformation process (for example, five thousand nuclei). In the second situation, there is no site saturation, and the nuclei appear as the older ones grow, so *I* is a rate (for example, five thousand nuclei/s).

When we generalize Equation 1, we have Equation 3:

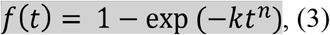

in which:

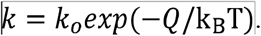

In a phase transformation process, Q represents the activation energy, k_B_ is the Boltzman constant, and T is the temperature.^12,14^ In practical cases, the values of *k* and *n* can assume different values, considering that the nuclei are not perfectly spherical, the nucleation rate is not constant, among other practical variables not predicted.

### Description of the cases of COVID-19 with the Johnson-Mehl-Avrami-Kolmogorov model

For the present paper, uncontaminated people will be the universe to be transformed from the uncontaminated to the contaminated state. Thus, for the application of the JMAK equation in cases of epidemics, we consider that:

- temperature *T* represents the degree of people circulating on the streets, in their neighborhoods, and making the pandemic grow at the contaminated border, without long-distance transport to form new nuclei;
- parameter *k* describes the expansion rate of COVID-19 cases over time — we consider *k* to be a constant at first approximation, since *T* is constant (represents the number of contaminated people advancing in the uncontaminated neighborhood at a constant rate);
- *f* is the fraction of people infected in a population;
- *n* represents the ability to create new nuclei of contaminated people and is associated to the transport of people in long-range means of transportation, such as by plane, subway, bus, and train.

The number of people recovered (immunized and killed) were not considered.

In our model, a person infected with COVID-19, when in a region not affected by the disease, can generate a nucleus (cluster 1) of infected people at a certain time, which we consider as the initial time (*t*=0) of the disease outbreak. In this situation, some people acquire the disease. At a certain moment, a person from that nucleus moves to another region where there is no infection by some form of public transport, and a second nucleus may appear (cluster 2). We consider that the clusters have different sizes because they were created at different times, and the contaminated nucleus grows over time, as shown in Figures 1A and 1B. If no measure to contain the disease outbreak is taken, other people infected with the disease create new nuclei in regions not yet affected by the virus. At a given time *t*_*x*_> 0, we will have other nuclei formed, as shown in Figure 1C.

**Figure 1.**
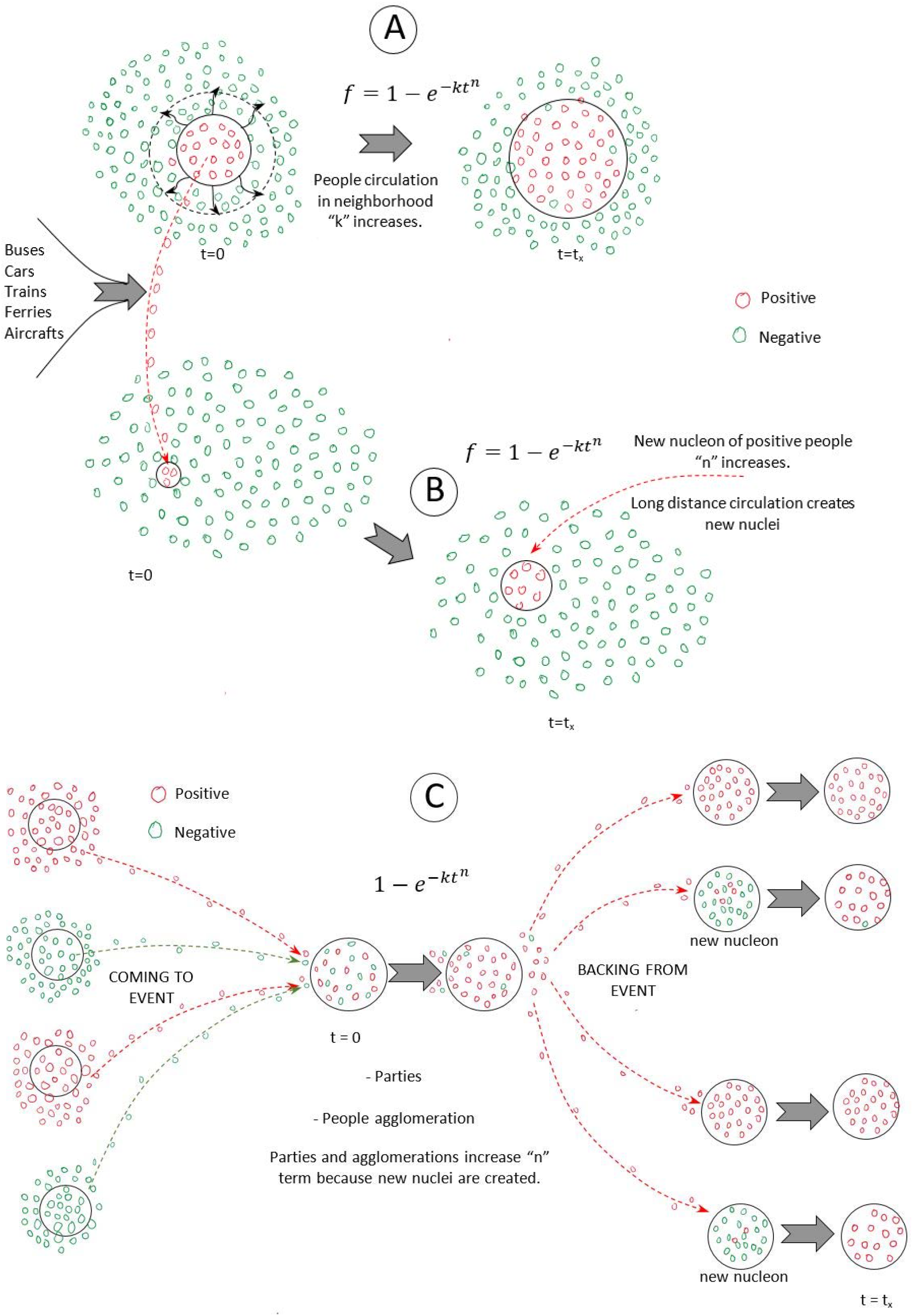
Nucleation model of COVID-19.

Depending on the types of events (parties, concerts, football matches, etc.) that cause crowds of people, there is a rapid growth of a nucleus and the creation of new clusters that are strongly dependent on the type of social mobility (plane, subway, bus, and train). In this description, the model does not allow us to measure the mobility parameter. However, the ease of mobility is related to parameter *n* of the JMAK function. If the value of *n* is equal to one, no new nuclei are created; but the higher its value, the more nuclei are generated. For n> 2 we have an indication that the disease spread is out of control.

### Determining the adjustment parameters

For obtaining the parameters *n* and *k*, we linearize Equation 3 in Equation 4:

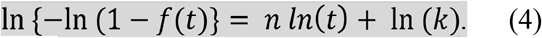

Then, we build the graph ln {−ln(1 − *f*(*t*)} *versus n ln* (*t*), as shown in Figure 2.

**Figure 2.**
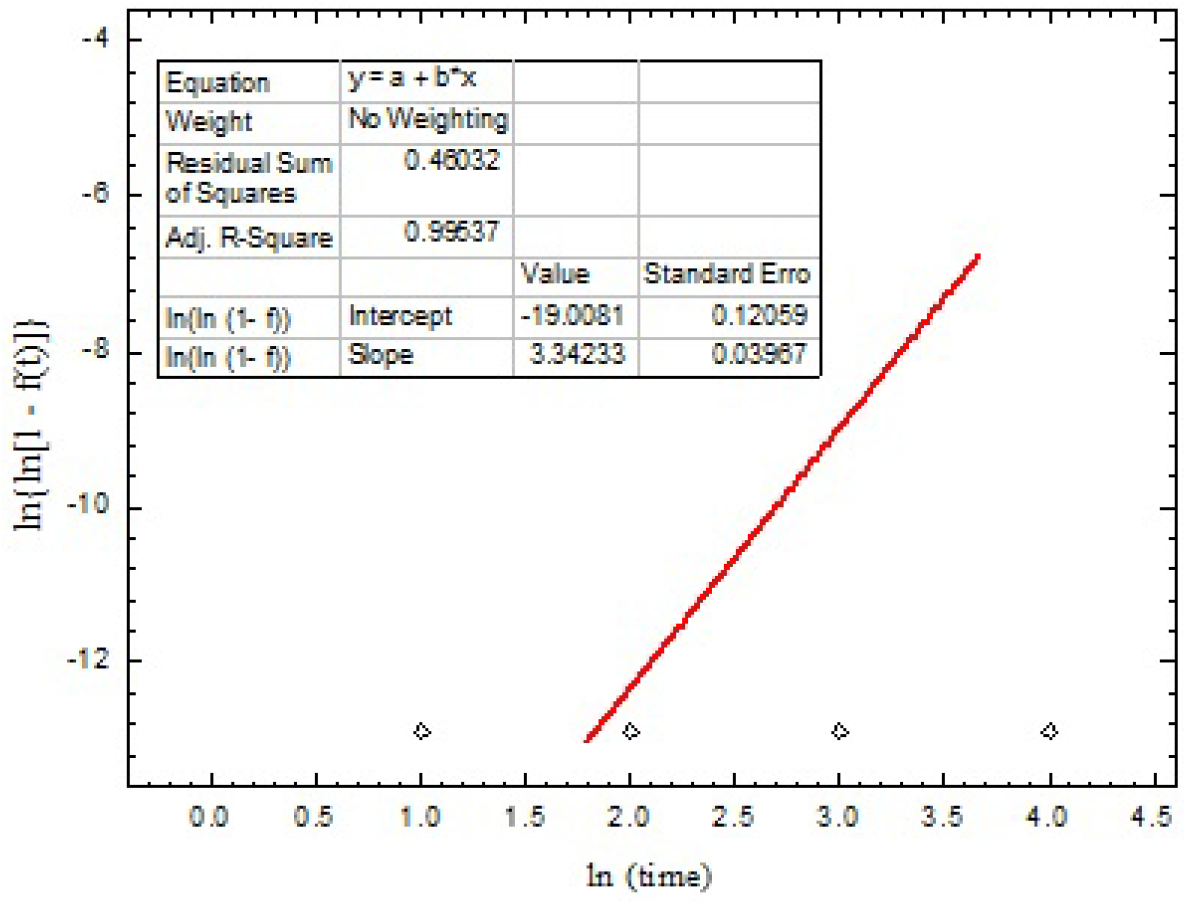
The line was adjusted in the interval between the 6^th^ and the 39^th^ day, with R^2^=0.99537.

We then use these adjustment parameters to find the number of total cases and the daily number of cases. For this purpose, knowing the total population of the analyzed region is needed.

## RESULTS

### Prediction of the number of people infected calculated with the Johnson-Mehl-Avrami-Kolmogorov function

#### Estimate with the percentage of infected people for Brazil

Countries that have already experienced the peak of the COVID-19 outbreak have different percentages of infected people, as described in Table 1. Thus, our insight is that the disease will affect between 1 and 3% of the Brazilian population.

**Table 1.** Number of confirmed cases of COVID-19 in some countries.

| Country | No. of infected people | Population | p% |
| --- | --- | --- | --- |
| Italy | 197,641 | 60,461,826 | 0.327 |
| Canada | 48,489 | 37,742,154 | 0.128 |
| South Korea | 11,018 | 56,263,639 | 0.020 |
| Australia | 7,019 | 25,811,863 | 0.027 |
| United Kingdom | 236,711 | 67,840,351 | 0.349 |
| Italy | 223,885 | 60,472,892 | 0.370 |
| United States | 1,471,967 | 330,753,490 | 0.445 |
| Spain | 274,367 | 46,752,506 | 0.587 |
| Brazil | 241,080 | 212,376,810 | 0.114 |
| Espírito Santo State | 5,013 | 4,018,650 | 0.125 |

Source: Wodermeter<sup>16</sup>.

The results of simulations are shown in Figure 3. Our estimate indicates that the peak of the disease will occur on June 29^th^ and will affect 29,402 infected people on that day. In addition, the disease’s nucleation rate is 4.5, which indicates that the disease is expanding very rapidly in Brazil.

**Figure 3.**
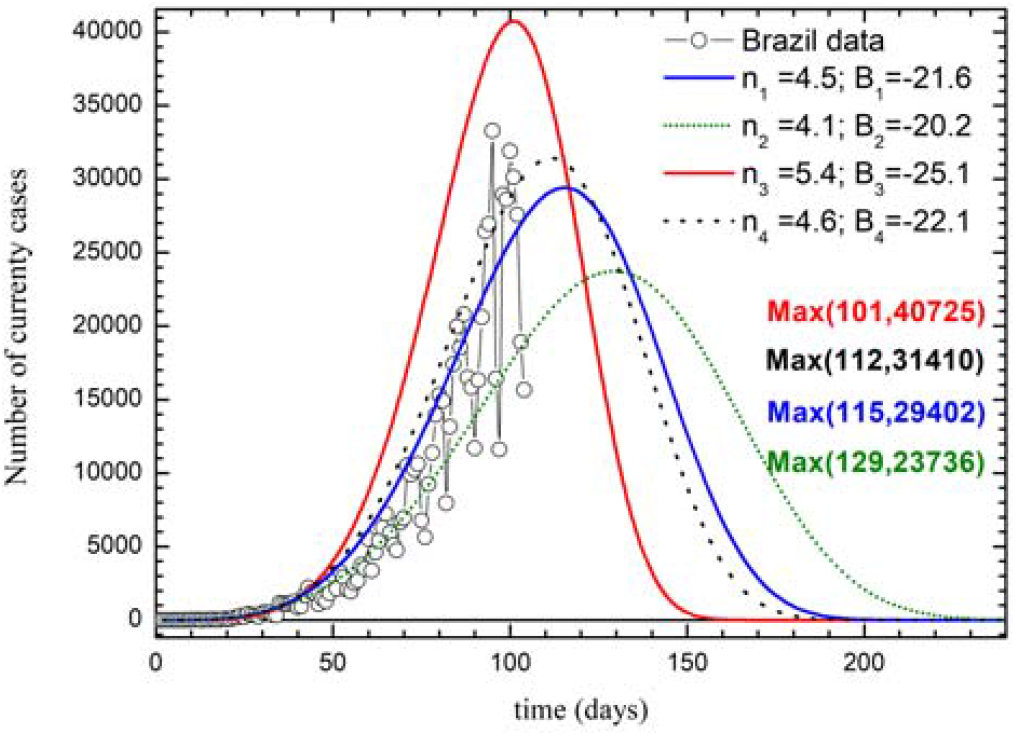
Estimate of people infected with COVID-19 considering 1% of the infected Brazilian population.

#### Prediction for Espírito Santo State

COVID-19 was first detected in Brazil in the eighth epidemiological week, with nine confirmed cases. Since the first one, health authorities have not carried out mass testing, suggesting the underreporting of the disease cases. The Brazilian Ministry of Health detected a 705% increase in admissions for severe acute respiratory syndrome (SARS) up to the 21^st^ epidemiological week of 2020, compared to the same period in 2019. We obtained this information on June 4^th^, on the Brazilian Ministry of Health website.

Regarding such data, Cintra and Nunes^10^ noted that, the number of hospitalizations for SARS-CoV-2 in Brazil was 12,260 in the 13^th^ epidemiological week, whereas the number was 1,123 in 2019. Given the average history between 2017 and 2019, 90% of hospitalizations are excessive cases that the researchers associated to COVID-19. Likewise, Bastos et al.^17^ observed a significant increase in hospitalizations for SARS when analyzing the historical average between the 9^th^ and 12^th^ epidemiological weeks of the decade of 2010. This reinforces our hypothesis that the Brazilian health system underreports COVID-19 cases.

To estimate the underreporting factor, we analyzed the data on deaths from COVID-19. The COVID Registral^18^ Panel website reports that there are six deaths from SARS in 2019 *versus* 62 deaths in 2020, an increase of approximately 1,000%. Then, we can define factor 10 as an underreporting correction. Therefore, if we correct the COVID-19 data by factor 10, we have the following evolutions in the number of cases, which can be seen in Figure 4A:

**Figura 4.**
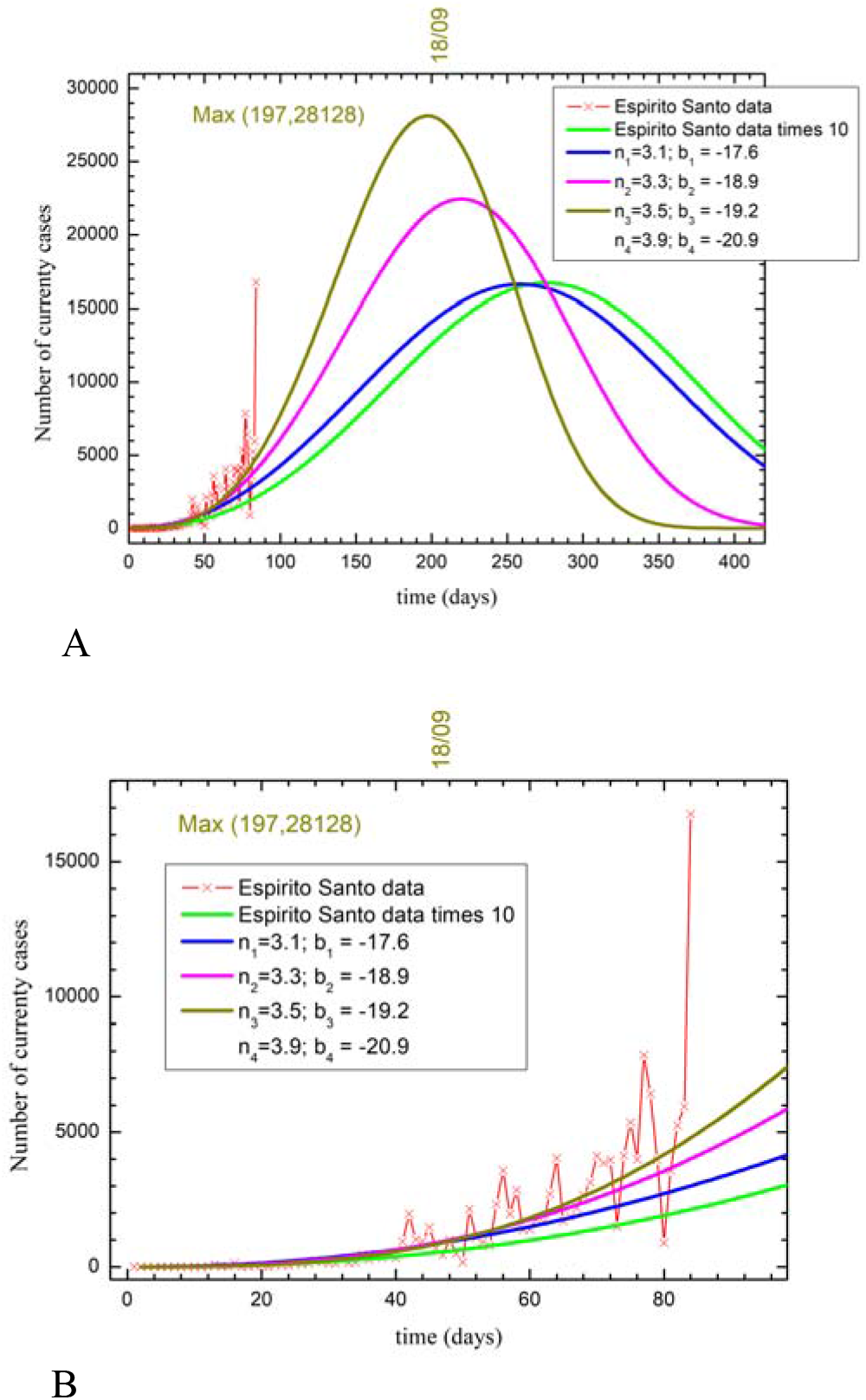
(A) Oficial data on COVID-19 corrected by the factor 10x. The closed square symbol represents the official data, and the cross symbol, the official data multiplied by 10. (B) Expansion of adjustments with the JMAK function.

As seen in the expansion (see Figure 4B), in the dark yellow curve (n_4_; b_4_), we will have the peak on day 197 (September 18^th^), with 28,128 people infected. In this epidemiological scenario, there will be 4.019 million people infected, which is a very bad estimate, because the total number of infected people is close to the total population of Espírito Santo State.

Considering 1% of the population infected by COVID-19 in Espírito Santo State, the adjustment parameters are *n*=3.8 and *b*=18, as shown in Figure 5. The difference between the official and simulated data agree. However, in the last epidemiological week, there was a big data variation, which can be explained by the number of people circulating during the week, compared to the weekend.

**Figure 5.**
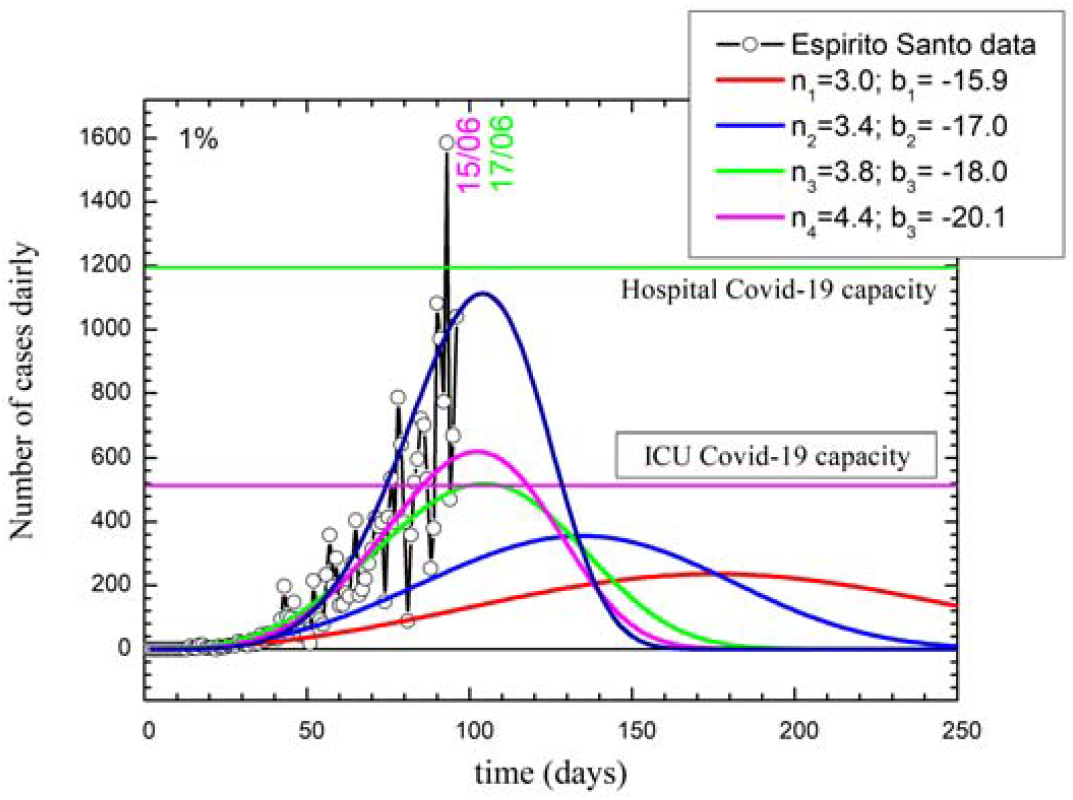
Estimate of people infected with COVID-19 considering 1% of the infected population of Espírito Santo State.

The peak of infection is estimated to occur on day 105 (June 19t^h^, 2020), with 519 people infected on this day. According to the COVID-19 Panel on Espírito Santo State, there are 490 beds in the intensive care unit (ICU) dedicated to COVID-19, of which 473 (60.03%) are occupied (see Table 2). In this epidemiological scenario, the state’s health system will not support the demand for beds.

**Tabela 2.**
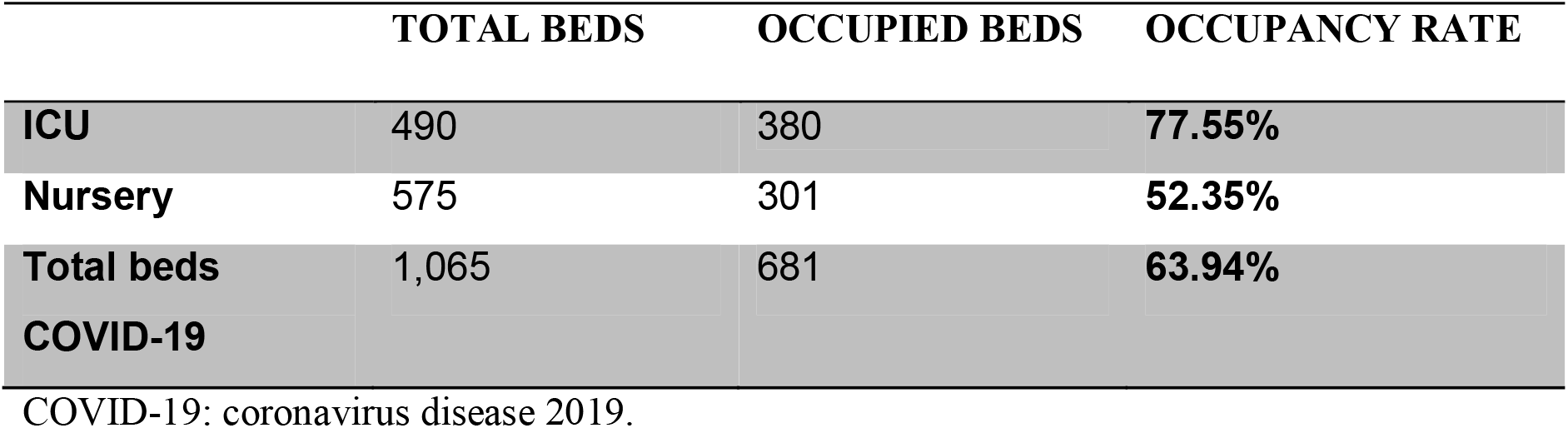
Beds in hospitals in Espírito Santo State. Update of beds in intensive care units (ICU) on May 25^th^, 2020.

#### Prediction for Pará State

Figure 6 shows the evolution and simulations for Pará State. Considering that 1% of the population is infected with COVID-19, the adjustment parameters that describe the evolution of daily cases are *n*=5.7 and *b*=24.9. Despite daily variations, the difference between official and simulated data is in good agreement.

**Figure 6.**
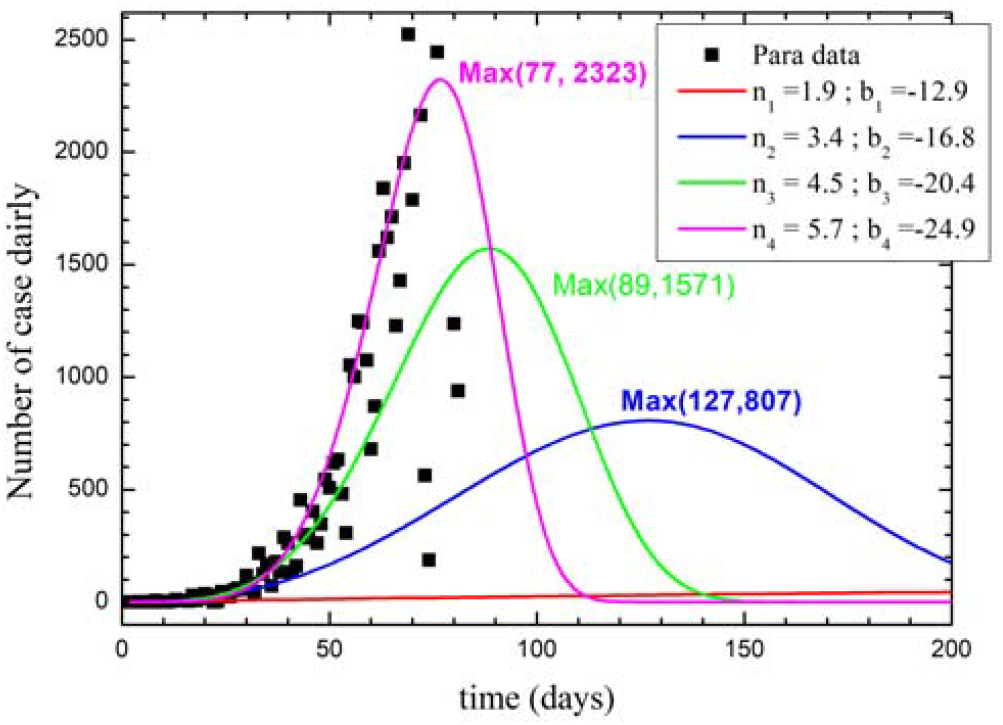
Estimate of people infected with COVID-19 considering 1% of the infected population of Pará State.

Parameter *n* found for Pará State is highlighted for having a higher value than that determined for Brazil (4.5). The first case registered in the state was on March 19^th^, and the state government implemented measures of social isolation. In Bélem City, the city hall decreed a lockdown on May 5^th^ for 20 days. However, there was a strong growth of the disease when compared to the rest of the country. Our result suggests that the mobility restriction measures did not work in Pará State.

## FINAL CONSIDERATIONS

In the present study, we analyze the official data of COVID-19 in Brazil and use the JMAK function to predict the disease evolution. The model indicated that the nucleation rate is in the order of four, which suggests that Brazilians are gathering without respecting measures of social distance and disease prevention. In our opinion, the political authorities were unable to control the spread of the disease in Brazil, considering that social mobility was partially interrupted by the federal and state governments. Summarizing the results, we have the estimate that 1% of the Brazilian population is infected:

- peak estimated to occur on June 29^th^;
- total number of infected people = 2.1 million.

As to Espírito Santo State, the estimate says that 1% of the state’s population is infected:

- peak estimated to occur on June 19^th^;
- total number of infected people = 40,186.

Estimating a scenario for Pará State, 1% of the population is infected:

- peak estimated to occur on June 3^rd^;
- total number of infected people = 86,029.

Given these scenarios, free movement of people is not recommended. In our opinion, the only possible alternative now is to keep social distancing measures, avoid gathering, and restrain access of people to public transportation.

## Data Availability

We searched for epidemiologic bulletins in all Brazilian local government websites and the Federal District that are summarized in .https://covid.saude.gov.br/

## AUTHORS’ CONTRIBUTION

Carlos Augusto Cardoso Passos: writing, collecting and analyzing data, building graphs, and interpreting data; Estéfano Aparecido Vieira: adequacy of JMAK theory and data interpretation; José André Lourenço: adequacy of JMAK theory and data interpretation; Jefferson Oliveira do Nascimento: data collection and interpretation.

